# A CT-based radiomics approach for CD8+ lymphocytes infiltration stratification in patients with non-small cell lung cancer

**DOI:** 10.1101/2024.05.23.24307791

**Authors:** Fadila Zerka, Mehdi Felfli, Charles M Voyton, Alexandre Thinnes, Sebastien Jacques, Yan Liu, Antoine Iannessi

**Affiliations:** Median Technologies, Valbonne, France; Centre Antoine Lacassagne, Nice, France

**Keywords:** CD8, TILs, NSCLC, Radiomics, CT

## Abstract

**Background:** Accurate prediction of tumor microenvironment is crucial for optimizing decision making throughout cancer treatment process. Current biopsy or surgical-based approaches to assess tumor microenvironment are limited by their invasiveness and tumor heterogeneity. The present study aimed to investigate the association of computed tomography radiomics features and CD8+ lymphocyte infiltration levels for patients with non-small cell lung cancer.

**Materials and Methods:** 283 patients with CT imaging and RNA-Seq data were collected from open-source data repositories. The study included three independent cohorts of non-small cell lung cancer patients, with one serving as the training set and the other two as external test sets. 1246 CT radiomics features were extracted. Three discriminative texture features were used to train the AI model.

**Results:** The model, trained on discriminative features, achieved a mean area under the curve AUC-ROC of 0.71(±0.17 std) on the training data. The AUC-ROC of the model on the two independent test sets is 0.67 (95% CI: 51%, 80%) on TCGA and 0.64 (95% CI: 51%, 74%) on LUNG3.

**Conclusion:** CT texture features can differentiate patients with high from low CD8+ lymphocyte infiltration levels. These features can non-invasively analyze the whole tumor and aid in the identification of patients that can respond to immunotherapy.

**Tweetable abstract:** Texture radiomics features on CT scans can aid in stratifying CD8+ lymphocyte infiltration levels for patients with NSCLC.

## Introduction

Lung cancer stands as the foremost cause of cancer-related mortality worldwide. Despite the progress in non-small cell lung cancer (NSCLC) treatments, surgical resection continues to be the primary choice for NSCLC patients. Nevertheless, surgery is not applicable for all patients, especially patients with metastasis outside the lungs (1). Treatment options for late-stage patients are diverse and prone to resistance and recurrence, hence, there is an urgent need for additional novel and compelling biomarkers that can aid in indicating the diagnosis and prognosis of lung cancer.

The progression of lung cancer involves both the tumor and the surrounding biological system, which is influenced by a combination of genetic factors and multiple cell interactions such as tumor host immune cells and stromal in case of metastasis (1). Immune evasion plays a pivotal role in the development of lung cancer, by hiding from or suppressing the immune response of the host cell. Therefore, the presence of tumor-infiltrating lymphocytes (TILs) in the tumor microenvironment serves as an indicator of the host immune response to tumor antigens (2). TILs are composed of diverse immune cell types such as natural killer (NK) cells, macrophages, and T cells including CD3+, CD4+, CD8+ lymphocytes, regulatory T cells, and others (3). Immunotherapeutic agents have been shown to improve survival in NSCLC (4,5). Depending on the distribution and frequency of immune cells, tumors can be classified into three main categories notably immune-inflamed, immune-desert, and immune-excluded (6). Tumors characterized by immune inflammation exhibit dense and active infiltration of CD8+ cells, indicating the presence of immune checkpoint inhibitors, like PD-L1, and an elevated mutational burden. Currently the gold standard for TILs quantification is the assessment of surgical or biopsy specimens via immunohistochemistry, transcriptomics, and cytometry. This approach, however, is limited by its invasiveness, limited reproducibility, and inability to reflect intra- and intertumoral heterogeneity. In addition to that many patients experience immune-related adverse effects. These effects can impact their quality of life, increases healthcare expenses, and in severe cases, can cause impairment or death (7). Therefore, noninvasive predictive biomarkers are needed to identify patients likely to benefit from immunotherapy, minimizing risks and enhancing treatment efficacy.

Quantitative imaging is a rapidly evolving field centered around the high throughput feature extraction from medical imaging, also known as radiomics features. Computed tomography (CT) image based radiomics is a non-invasive approach, allowing comprehensive analysis of the entire tumoral tissue, microenvironment, and its surroundings. Therefore, radiomics features facilitate the characterization of tumor spatial heterogeneity and enable the longitudinal evaluation of disease progression, evolution, and response to treatment.

Our study aims to investigate the ability of radiomics features alone to stratify patients based on CD8+ lymphocyte infiltration levels and identify relevant radiomics features associated with these levels.

## Material and methods

### Data

Three publicly available datasets containing CT images and gene expression data (RNA-Seq or Micro-Array) for patients with non-small cell lung cancer (NSCLC) were used to train and test the model. Only patients with both CT imaging and RNA-Seq (N=273) were eligible for further analysis. A flow chart describing the data flow of the study is shown in Fig 1.

**Fig 1.**
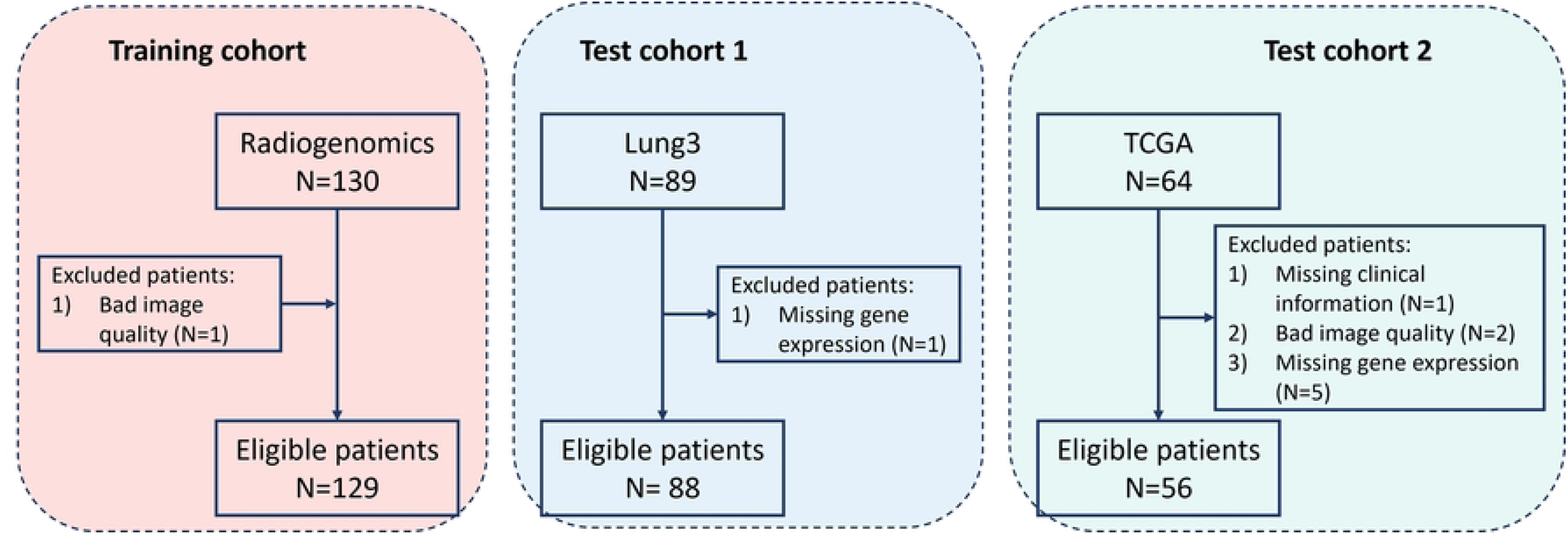
data flow

One dataset (referred to as Radiogenomics) was used for model training(8–10). The two other datasets were used for model testing. The first test set is radiomics-genomics, referred to as Lung3(11,12). Both were collected from The Cancer Imaging Archive (TCIA, http://cancerimagingarchive.net/) (13). The second test set consisted of two subsets from The Cancer Genome Atlas (TCGA), which included both squamous cell carcinoma and adenocarcinoma cases, namely TCGA-LUSC dataset and TCGA-LUAD respectively (14,15).

Among the 273 patients, 50 identified with multiple tumors (Radiogenomics N=31, Lung3 N=6, TCGA N=13). Only the tumor corresponding to the biopsied tumor description was retained for each patient, while any additional tumors were excluded.

### Immune microenvironment estimation

The TILs data was collected directly from the TIMER platform which calculated the immune profiles of each of the patients in the TCGA library. The TILs data for Radiogenomics and Lung3 datasets were not available. However, the RNA sequencing data was available and were collected from the Gene Expression Omnibus (GEO) (16,17). For Radiogenomics patients we calculated the immune profiles using the TIMER software platform from the raw data (18). We collected the RNA sequencing data for LUNG3 dataset, normalized using Robust Microarray Analysis (RMA) with default parameters. The normalized RNA sequencing data is then used to extract gene expression matrix using Bioconductor package (V3.18). Furthermore, TME composition was estimated using deconvolution methods(Immunedeconv R package) (V2.1) followed by tumor infiltration subtypes estimation. Distributions of CD8+ subtype are depicted in Fig 2.

**Fig 2.**
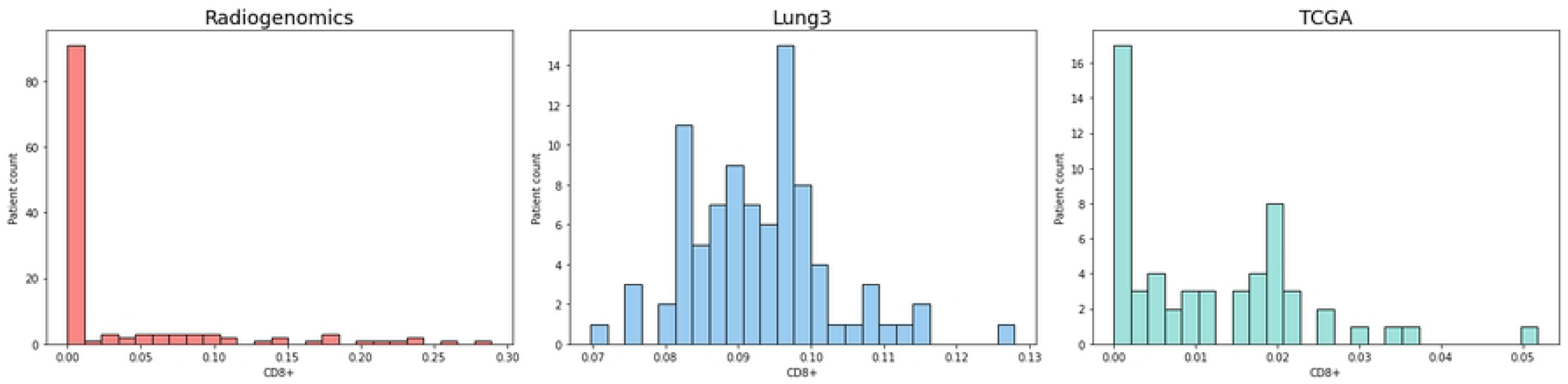
Distribution of CD8+ subtype of the Radiogenomics, Lung3, LUAD, and LUSC cohorts

### Binary outcome estimation

We computed the mean CD8+ levels for each dataset (Radiogenomics, Lung3, and TCGA) and utilized this average as a cut-off value to stratify patients into two groups: high CD8+ and low CD8+. As illustrated in Fig 2, the distribution of CD8+ levels varies among the different datasets. Consequently, we applied a distinct cut-off value for each set instead of a universal cut-off for all datasets combined. A high CD8+ expression was determined if it exceeded the cut-off point specific to the dataset in question; otherwise, it was considered low. The patient distribution for each group is detailed in Table 1.

**Table 1.**
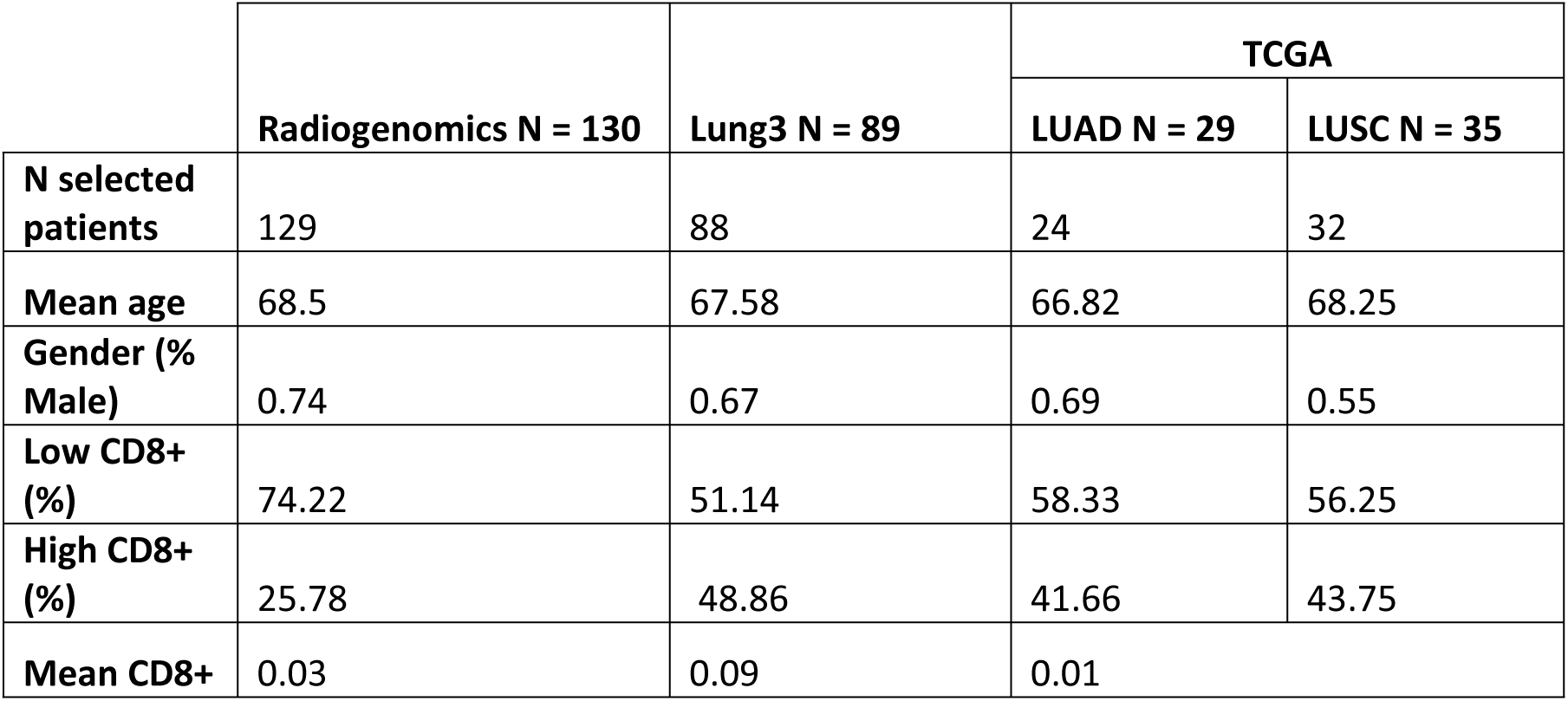
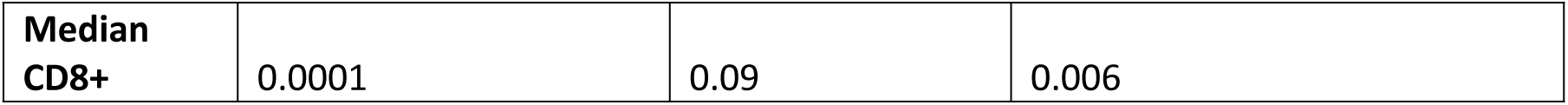
patient characteristics.

|  |  |  | TCGA |  |
| --- | --- | --- | --- | --- |
|  | Radiogenomics N = 130 | Lung3 N = 89 | LUAD N = 29 | LUSC N = 35 |
| **N selected patients** | 129 | 88 | 24 | 32 |
| **Mean age** | 68.5 | 67.58 | 66.82 | 68.25 |
| **Gender (% Male)** | 0.74 | 0.67 | 0.69 | 0.55 |
| **Low CD8+ (%)** | 74.22 | 51.14 | 58.33 | 56.25 |
| **High CD8+ (%)** | 25.78 | 48.86 | 41.66 | 43.75 |
| **Mean CD8+** | 0.03 | 0.09 | 0.01 |  |

|  |  |  |  |
| --- | --- | --- | --- |
| **Median**<br>**CD8+** | 0.0001 | 0.09 | 0.006 |

### Radiomics feature extraction

Radiomics refers to the extraction of large amounts of quantitative features from medical images, allowing for a thorough quantification and description of tumor phenotypes. The underlying concept of radiomics is that there are unique quantifiable features in medical imaging that can shed vital insights into tumor physiology, which could be leveraged to improve cancer diagnosis and prognosis.

Lesion delineation is required prior extracting the radiomics features. An in-house semi-automatic tool was used to segment lung lesions. The segmentations were reviewed and if needed edited by an experienced radiologist with 20+ years of experience. In this study, we did not implement any volume cut-off criteria for patient inclusion. The same software was used to convert DICOM objects to nifty arrays to be used for further processing. We employed the open-source PyRadiomics library (V3.1) for the extraction of radiomic features. Prior radiomics feature extraction all CT images were resampled to an isotropic grid of 1×1×1 mm^3^ using B-spline interpolation to consistently calculate the three-dimensional features. A fixed bin width of 25 Hounsfield units was used for image intensity discretization, therefore reducing noise and computational load. A multi-scale wavelet filter that computes eight decompositions (HHH, HHL, HLH, HLL, LHH, LHL, LLH, LLL) per level and Laplacian of Gaussian (LoG) filter (σ = 1.0, 2.0, 3.0, 4.0, 5.0mm) were applied to the images to extract filter features. For each lesion, 1246 radiomics features were extracted. The extracted features comprised 102 original features (18 first-order statistics, 70 texture (or otherwise known as second-order statistics), and 14 shape features) and 1144 filtered features (704 wavelet and 440 LoG features). The radiomics process applied in this study is depicted in Fig 3.

**Fig 3.**
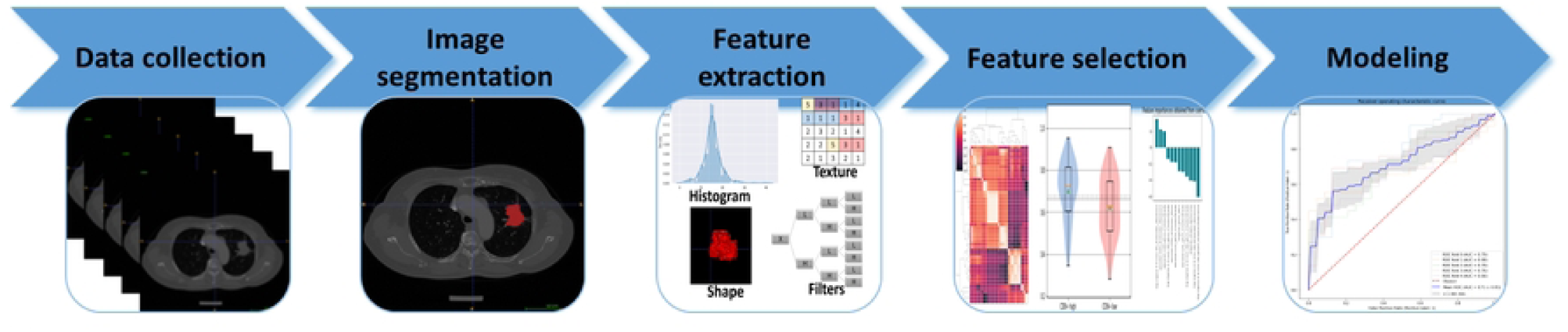
Schematic representation of the radiomics workflow: **Data collection:** CT and RNA-Seq data for patients with NSCLC collected from opensource repositories. **Image segmentation:** Tumors segmented using a semi-automatic tool. **Feature extraction:** Radiomics features were extracted from the segmented tumors. **Feature selection:** The robust radiomics features were selected, univariate analysis applied to the remaining features to test their ability to stratify patients with high CD8+ vs low CD8+, then feature importance analysis were applied to keep only important features for the model. **Modeling:** The discriminative radiomics features were selected and used to train the AI model and the performances were validated on the test sets.

### Statistical analysis

#### Univariate analysis

Before evaluating the associations between the radiomics features, we initially reduced the feature space, aiming to eliminate any redundant features. Features with near zero variance were first discarded. Then the highly correlated features with Pearson correlation coefficient above 0.8 and features with linear combinations between features were eliminated from further analysis. Then, patients were split into two groups (high CD8+/low CD8+). The normality of each feature from the remaining features was assessed using Shapiro-Wilk test. A Student’s t-test was used to compare groups with normal distributions, while the Wilcoxon rank sum test was used to compare features with non-normal distributions. A p-value under 0.05 was considered statistically significant. The features with significant discrimination of CD8+ patient groups were used for further analysis.

#### Feature importance

Hundred times repeated 10-fold cross validation was used to fit a logistic regression model. A different random estate was used at each iteration to ensure the uniqueness of the data subsets used to train and validate the models. Feature regression coefficient was saved at each iteration, and compared to the rest after training was over. The features with positive coefficients at every iteration were saved to train the final model.

#### Multivariate analysis

A logistic regression model (LR) was performed in the training data using a ten-fold cross validation to classify CD8+ cell infiltration level (high vs low). Model parameters were optimized prior training using the python GridSearchCV pipeline. All features were standardized before model training. Additionally, to correct class imbalance, and reduce overfitting risk to the most representative class, radiomics features were randomly oversampled before modelling. Model performance was validated in test data from TCGA and LUNG3 datasets. Here, the area under the receiver operating characteristic curve (AUC) was used to assess model performance in discriminating between high CD8+ and low CD8+ cell infiltration levels in patients with NSCLC. All statistical analysis were performed in Python V3.9.

## Results

### Data characteristics

The patient demographic characteristics are detailed in Table 1. A total number of 129 patients were used for model development 88, and 56 patients from LUNG3 and TCGA were respectively used for independent model testing.

### CD8+ infiltration prediction

The univariate and features importance analysis yielded three discriminative texture type radiomics features with significant differences between the two groups (high and low CD8+). The discriminative features consisted of 1) log-sigma-4-0-mm-3D_firstorder_Skewness, 2) wavelet-LHH_firstorder_Skewness, and 3) wavelet-LHH_glcm_MCC, with p-val = 0.028239, p-val = 0.026691, p-val = 0.016508 respectively. We conducted a univariate analysis of the discriminative features on both external test sets to evaluate the distinction between the two groups, respectively. The results are presented in violin box plots and quantile plots, as shown in Fig 4.

**Fig 4.**
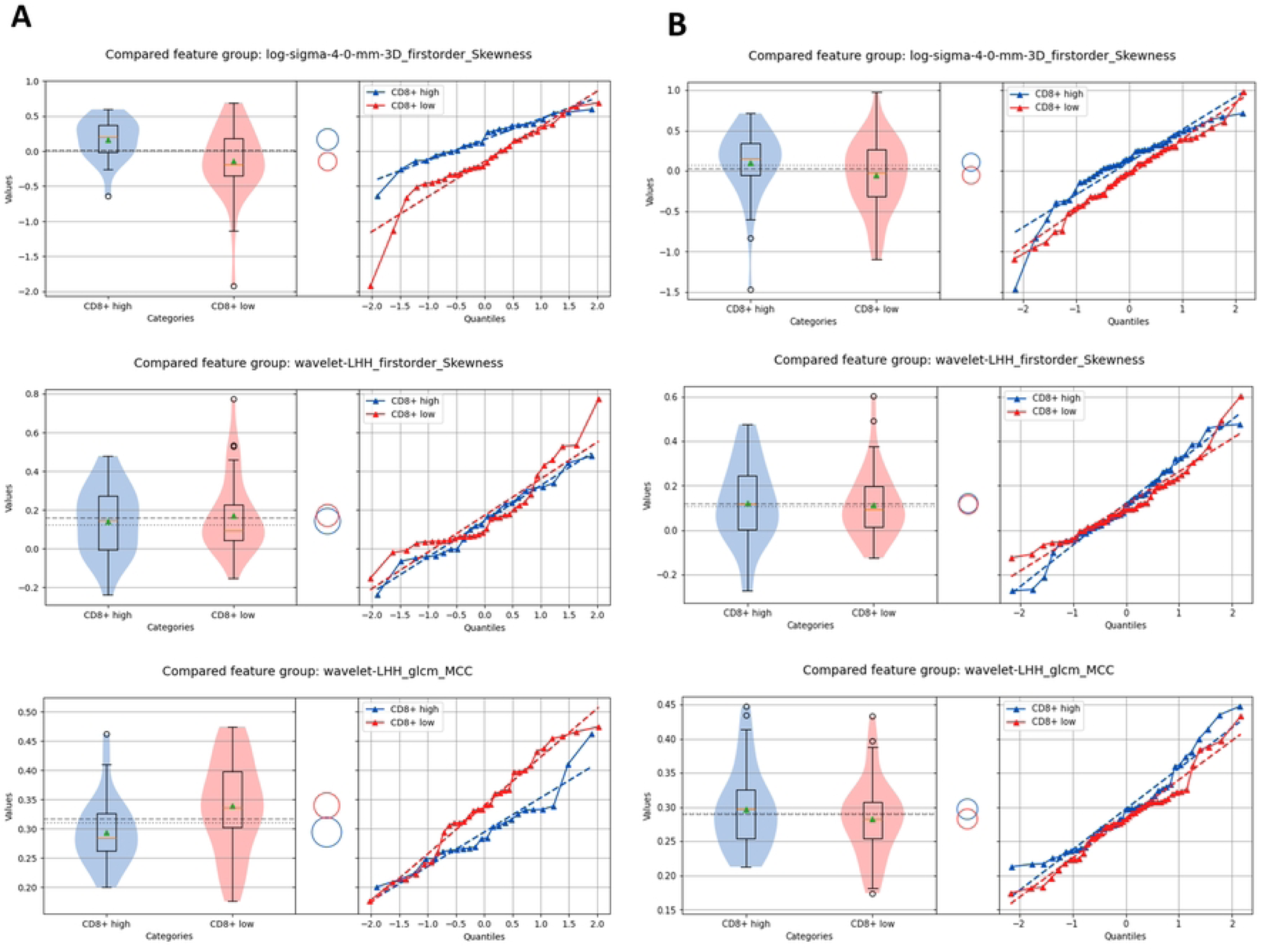
Violin box plots and quantile plots comparing the discriminative features for patients with high and low CD8+ infiltration levels; (A) TCGA test set, (B) LUNG3 test set.

Given that the imaging biomarker profile was able to discriminate CD8+ infiltration levels, we trained a logistic regression model to evaluate whether this imaging biomarker retained predictive ability for CD8+ patient groups. The mean AUC of the radiomics in the training set was 0.71(±0.17 std), 0.67 (95% CI: 51%, 80%) on the TCGA independent test set, and 0.64 (95% CI: 51%, 74%) on the LUNG3 test set. The Receiver operating characteristic curve ROC curve for the training data is depicted in Fig 5A. Whereas the ROC curves for the independent test sets are shown in Fig 5B.

**Fig 5.**
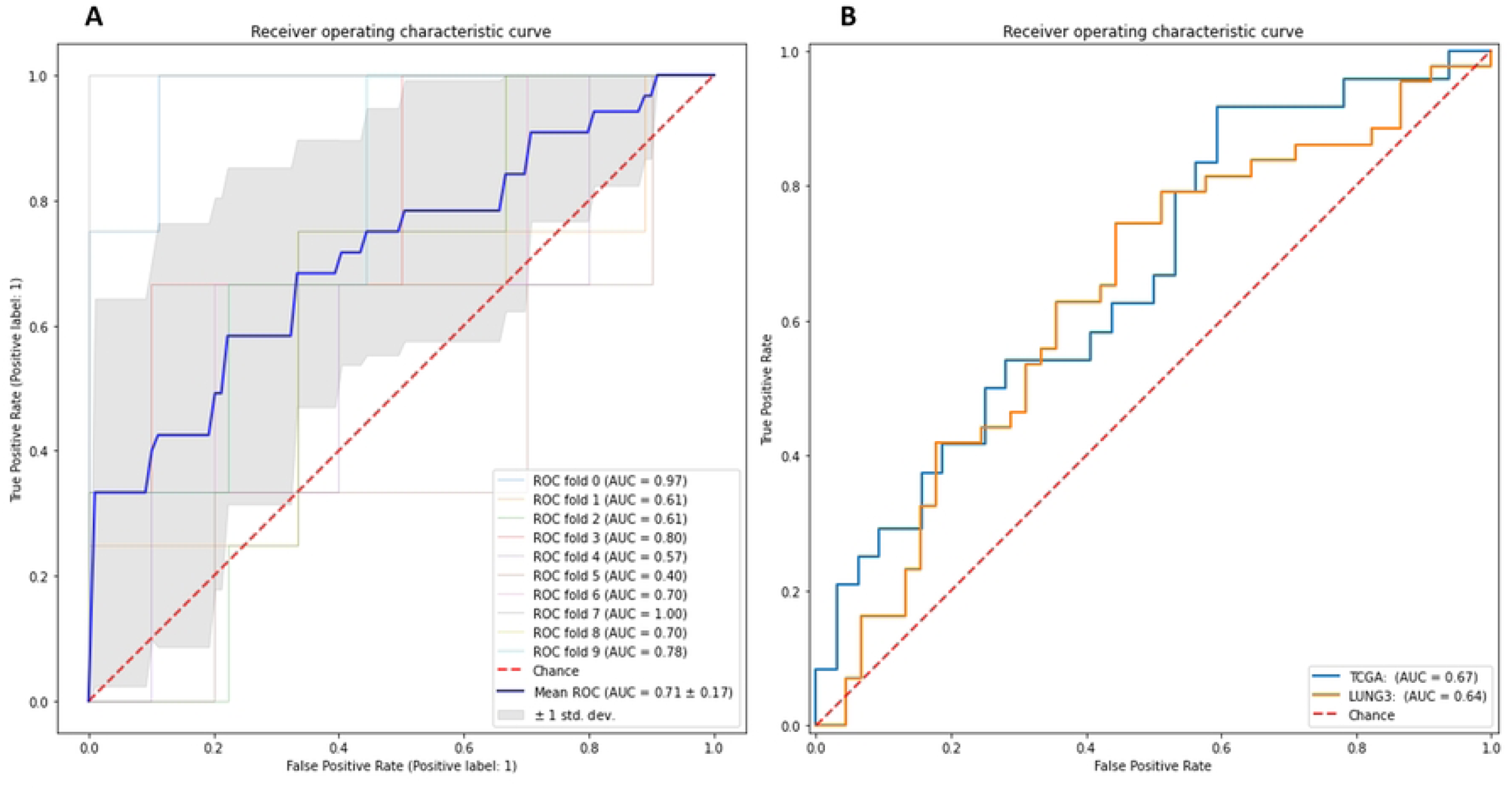
(A) AUC ROC curves of the 10-fold cross validation radiomics-based models; (B) AUC ROC curves for CD8+ infiltration high and low patient prediction in the independent test sets.

We compiled Kaplan-Meier curves for overall survival for Radiogenomics and TCGA datasets. survival curves are depicted in Fig 6. Within each cohort the patients were stratified into two groups (high CD8+ and low CD8+). LUNG3 dataset did not have event timepoint, therefore we did not conduct survival analysis for this dataset.

**Fig 6.**
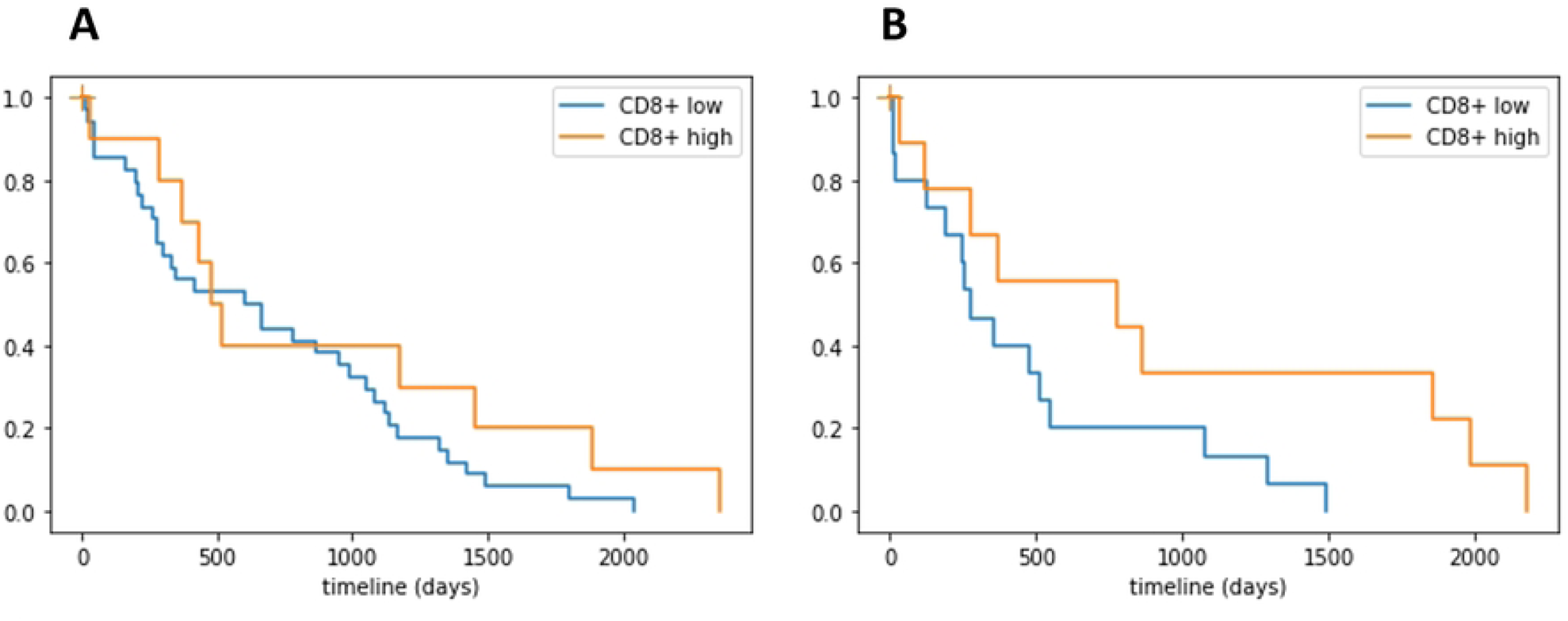
Kaplan–Meier curves demonstrating performance of the radiomic signature on (A) the training cohort and (B) the TCGA cohort

## Discussions

Cancer treatment is increasingly relying on immunotherapy (19). Tumor microenvironment, in particular, tumor-infiltrating immune cells analysis have previously unveiled connections between immunogenic markers and treatment response factors, such as survival or tumor growth, and other clinical outcomes (20–22). Despite the extensive research in this domain, the relationship between tumors and their microenvironment is not yet fully unraveled. Moreover, the prevailing studies often rely solely on immunohistochemistry-based analyses to assess tumor-infiltrating immune cells. The limitations of these approaches are evident when it comes to capturing tumor heterogeneity, mainly due to their reliance on small biopsy samples or surgical resection. Consequently, the absence of a valid noninvasive method capturing the entirety of the tumor microenvironment stresses the urgent requirement for the development of new biomarkers to predict and characterize the tumor microenvironment accurately. While CT image based radiomics is a non-invasive approach, allowing comprehensive analysis of the entire tumoral tissue, it stands as a potential alternative in addressing these challenges (11). Nevertheless, the literature linking the physiological tumor context, like radiomics, to immune response assessment is limited due to data heterogeneity and lack of reproducibility (23).

CD8+ is predominantly expressed on cytotoxic T lymphocytes (CTLs), which eliminates tumor cells through the release of perforin and the mediation of immune suppression (24,25). Therefore, they stand as the most dominant effect in the anticancer immune response and constitute the foundation of current successful cancer immunotherapies (26). CD8+ infiltration in tumor tissue and the surrounding stroma measured via pathology and cytology has been shown to be a strong predictor of ICI response (23,27). Accurate and fast prediction CD8+ infiltration levels could significantly enhance clinicians’ decision-making process, mainly by facilitating reaching personalized therapeutic consensus, since patients with high CD8+ infiltration levels are most likely to respond well to immunotherapy.

In this study, we examined the ability radiomics features extracted from CT images to discriminate CD8+ infiltration levels in patients with NSCLC. The data originated from three opensource cohorts. A total of 1246 radiomics features were extracted from each lesion. We identified three first order and textural features that were able to discriminate CD8+ infiltration levels in both the training and independent test cohorts. Our findings indicate that radiomics texture features can play a significant role in capturing tumor immune phenotypes. This suggests that radiomics features have the capacity to unveil a more complex range of phenotypic characteristics compared to conventional clinical factors.

Several other studies have shown that CD8+ infiltration levels significantly correlated with CT texture features for patients with NSCLC. Zhou et.al., reported that texture heterogeneity (NGLDM contrast) had a strong correlation with CD8+ infiltration levels (28). Another study reported that texture radiomics features (First-order, GLCM, GLRLM, and GLDM) correlate with not only CD8 infiltration levels but also with CD3 and PD-L1 (29). Similarly, Chen et.al., reported that six radiomics features (first order and five wavelet flittered texture features) significantly correlated with CD8 TILs (30). Min et.al., demonstrated that homogeneity and high grey-level values (GLCM and GLDM) correlate with CD8 and CD103 markers (31). However, reproducing the results reported throughout all those studies is not trivial due to the sensitivity of radiomics features to acquisition parameters and scanner types. Despite the heterogeneity of the data we used in our study, our findings validated the results presented in the previously published articles. This validation pertains specifically to the feature group, in this case, first order and texture features, rather than extending to individual feature sets. All the datasets used in the current study are publicly available, easing reproducibility of the results reported.

Despite the increased exploration of CT radiomics in clinical practice, substantial standardization challenges persist in the form of variations in study design and analysis methods. In addition to data variance is susceptibility to multiple parameters, including image acquisitions, reconstructions, and segmentations. Another limitation we observed when conducting this study is the variety of the TILs quantification methods used across the datasets, for example Illumina Hiseq 2500 (RNA-Seq) was used in Radiogenomics dataset, whereas Affymetrix FS450 (micro-array) for LUNG3 dataset. This variation might begin to explain the wide difference in the generated data distributions (Fig 2). Moreover, there is considerable variation in preparation protocols and histopathological assessment of specimens among different institutions. On top of this, the challenge of inter-tumor heterogeneity exists, as histological and cellular heterogeneity in lung cancer is also well documented with tumors having more than one type of differentiation (32). In the other hand clear stratification guidelines for tumor-infiltrating immune cells into high and low groups are not yet established. Consequently, we utilized the mean value to categorize our data into high or low levels of CD8+. Both mean and median counts are often used for categorizing high or low expression levels of tumor-infiltrating lymphocytes (TILs), yet neither is an optimal approach for individual decision-making, as these are specific population-based metrics (33).

In consequence, it is essential to implement standardized data acquisition and post processing protocols for both radiomics analysis and TILs expression to enhance the accuracy of prediction and facilitate reproducibility of the results. Additionally, the small size and heterogeneity of training data affected the generalizability of the model. This is in part because the distribution of the training data was different from the independent test sets (Appendices Fig S1, S2, and S3), and in the other hand, the acquisition and reconstruction parameters are different in all the cohorts making each subset poorly represented and therefore affected the reproducibility of the radiomics features. Increasing the sample size in addition to CT intensity harmonization using more advanced techniques such as generative adversarial networks (GAN) (34), may significantly help mitigate this limitation. In the future, defining clear stratification guidelines for TILs subtypes may greatly affect AI model development and ease reproducibility of the results.

## Conclusions

Texture radiomics features on CT scans can aid in distinguishing between patients with CD8+ lymphocyte infiltration levels below the mean (low) and those above the mean (high). Even if these features offer promise as surrogate predictors for patient responses to immunotherapy and aiding clinical decision-making, the lack of a consensus threshold for defining low and high infiltration levels hampers result generalizability and practical applicability.

## Data Availability

Raw all imaging data is publicly available from the http://cancerimagingarchive.net/ as well as genomic data of Lung3 and Radiogenomics radiogenomics datasets. Genomics data of LUSC and LUAD is available under TIMER platform (http://timer.cistrome.org/). All relevant results data are within the manuscript and its Supporting Information files.

https://wiki.cancerimagingarchive.net/display/Public/NSCLC-Radiomics-Genomics

https://www.cancerimagingarchive.net/collection/nsclc-radiogenomics/

https://www.cancerimagingarchive.net/collection/tcga-luad/

https://www.cancerimagingarchive.net/collection/tcga-lusc/

## Acknowledgments

The authors thank Dr. Y. Liu for editing the manuscript.

## References

1. Popper HH. Progression and metastasis of lung cancer. Cancer Metastasis Rev. 2016;35:75–91.

2. Hendry S, Salgado R, Gevaert T, Russell PA, John T, Thapa B, et al. Assessing Tumor-infiltrating Lymphocytes in Solid Tumors: A Practical Review for Pathologists and Proposal for a Standardized Method From the International Immunooncology Biomarkers Working Group: Part 1: Assessing the Host Immune Response, TILs in Invasive Breast Carcinoma and Ductal Carcinoma In Situ, Metastatic Tumor Deposits and Areas for Further Research. Adv Anat Pathol. 2017 Sep;24(5):235–51.

3. Chen B, Li H, Liu C, Xiang X, Wang S, Wu A, et al. Prognostic value of the common tumour-infiltrating lymphocyte subtypes for patients with non-small cell lung cancer: A meta-analysis. Lee MC, editor. PLOS ONE. 2020 Nov 10;15(11):e0242173.

4. Zhou GW, Xiong Y, Chen S, Xia F, Li Q, Hu J. Anti-PD-1/PD-L1 antibody therapy for pretreated advanced nonsmall-cell lung cancer: A meta-analysis of randomized clinical trials. Medicine (Baltimore). 2016 Aug;95(35):e4611.

5. Tanvetyanon T, Creelan BC, Antonia SJ. The safety and efficacy of nivolumab in advanced (metastatic) non-small cell lung cancer. Expert Rev Anticancer Ther. 2016 Sep;16(9):903–10.

6. Tiwari A, Oravecz T, Dillon LA, Italiano A, Audoly L, Fridman WH, et al. Towards a consensus definition of immune exclusion in cancer. Front Immunol [Internet]. 2023 [cited 2024 Jan 31];14. Available from: https://www.frontiersin.org/articles/10.3389/fimmu.2023.1084887

7. Mamdani H, Matosevic S, Khalid AB, Durm G, Jalal SI. Immunotherapy in Lung Cancer: Current Landscape and Future Directions. Front Immunol. 2022 Feb 9;13:823618.

8. Bakr S, Gevaert O, Echegaray S, Ayers K, Zhou M, Shafiq M, et al. Data for NSCLC Radiogenomics Collection [Internet]. The Cancer Imaging Archive; 2017 [cited 2023 Oct 26]. Available from: https://wiki.cancerimagingarchive.net/x/W4G1AQ

9. Bakr S, Gevaert O, Echegaray S, Ayers K, Zhou M, Shafiq M, et al. A radiogenomic dataset of non-small cell lung cancer. Sci Data. 2018 Oct 16;5(1):180202.

10. Gevaert O, Xu J, Hoang CD, Leung AN, Xu Y, Quon A, et al. Non–Small Cell Lung Cancer: Identifying Prognostic Imaging Biomarkers by Leveraging Public Gene Expression Microarray Data—Methods and Preliminary Results. Radiology. 2012 Aug;264(2):387–96.

11. Aerts HJWL, Velazquez ER, Leijenaar RTH, Parmar C, Grossmann P, Carvalho S, et al. Decoding tumour phenotype by noninvasive imaging using a quantitative radiomics approach. Nat Commun. 2014 Sep 22;5(1):4006.

12. Aerts HJWL, Rios Velazquez E, Leijenaar RTH, Parmar C, Grossmann P, Carvalho S, et al. Data From NSCLC-Radiomics-Genomics [Internet]. The Cancer Imaging Archive; 2015 [cited 2023 Oct 26]. Available from: https://wiki.cancerimagingarchive.net/x/GAL1

13. Clark K, Vendt B, Smith K, Freymann J, Kirby J, Koppel P, et al. The Cancer Imaging Archive (TCIA): Maintaining and Operating a Public Information Repository. J Digit Imaging. 2013 Dec;26(6):1045– 57.

14. Hammerman PS, Lawrence MS, Voet D, Jing R, Cibulskis K, Sivachenko A, et al. Comprehensive genomic characterization of squamous cell lung cancers. Nature. 2012 Sep;489(7417):519–25.

15. Collisson EA, Campbell JD, Brooks AN, Berger AH, Lee W, Chmielecki J, et al. Comprehensive molecular profiling of lung adenocarcinoma. Nature. 2014 Jul;511(7511):543–50.

16. GEO Accession viewer - Radiogenomics [Internet]. [cited 2024 Feb 22]. Available from: https://www.ncbi.nlm.nih.gov/geo/query/acc.cgi?acc=GSE103584

17. GEO Accession viewer - LUNG3 [Internet]. [cited 2024 Feb 22]. Available from: https://www.ncbi.nlm.nih.gov/geo/query/acc.cgi?acc=GSM1416530

18. Li T, Fu J, Zeng Z, Cohen D, Li J, Chen Q, et al. TIMER2.0 for analysis of tumor-infiltrating immune cells. Nucleic Acids Res. 2020 Jul 2;48(W1):W509–14.

19. Thorsson V, Gibbs DL, Brown SD, Wolf D, Bortone DS, Ou Yang TH, et al. The Immune Landscape of Cancer. Immunity. 2018 Apr;48(4):812–830.e14.

20. Hu Z, Gu X, Zhong R, Zhong H. Tumor-infiltrating CD45RO+ memory cells correlate with favorable prognosis in patients with lung adenocarcinoma. J Thorac Dis. 2018 Apr;10(4):2089–99.

21. Hugo W, Zaretsky JM, Sun L, Song C, Moreno BH, Hu-Lieskovan S, et al. Genomic and Transcriptomic Features of Response to Anti-PD-1 Therapy in Metastatic Melanoma. Cell. 2016 Mar 3;165(1):35.

22. Herbst RS, Soria JC, Kowanetz M, Fine GD, Hamid O, Gordon MS, et al. Predictive correlates of response to the anti-PD-L1 antibody MPDL3280A in cancer patients. Nature. 2014 Nov 27;515(7528):563–7.

23. Ramlee S, Hulse D, Bernatowicz K, Pérez-López R, Sala E, Aloj L. Radiomic Signatures Associated with CD8+ Tumour-Infiltrating Lymphocytes: A Systematic Review and Quality Assessment Study. Cancers. 2022 Jul 27;14(15):3656.

24. Ostrand-Rosenberg S, Sinha P, Beury DW, Clements VK. Cross-talk between myeloid-derived suppressor cells (MDSC), macrophages, and dendritic cells enhances tumor-induced immune suppression. Semin Cancer Biol. 2012 Aug;22(4):275–81.

25. Hatogai K, Kitano S, Fujii S, Kojima T, Daiko H, Nomura S, et al. Comprehensive immunohistochemical analysis of tumor microenvironment immune status in esophageal squamous cell carcinoma. Oncotarget. 2016 Jun 15;7(30):47252–64.

26. Raskov H, Orhan A, Christensen JP, Gögenur I. Cytotoxic CD8+ T cells in cancer and cancer immunotherapy. Br J Cancer. 2021 Jan;124(2):359–67.

27. Li F, Li C, Cai X, Xie Z, Zhou L, Cheng B, et al. The association between CD8+ tumor-infiltrating lymphocytes and the clinical outcome of cancer immunotherapy: A systematic review and meta-analysis. eClinicalMedicine. 2021 Nov;41:101134.

28. Zhou J, Zou S, Kuang D, Yan J, Zhao J, Zhu X. A Novel Approach Using FDG-PET/CT-Based Radiomics to Assess Tumor Immune Phenotypes in Patients With Non-Small Cell Lung Cancer. Front Oncol [Internet]. 2021 [cited 2023 Nov 13];11. Available from: https://www.frontiersin.org/articles/10.3389/fonc.2021.769272

29. Mazzaschi G, Milanese G, Pagano P, Madeddu D, Gnetti L, Trentini F, et al. Integrated CT imaging and tissue immune features disclose a radio-immune signature with high prognostic impact on surgically resected NSCLC. Lung Cancer. 2020 Jun;144:30–9.

30. Chen L, Chen L, Ni H, Shen L, Wei J, Xia Y, et al. Prediction of CD3 T cells and CD8 T cells expression levels in non-small cell lung cancer based on radiomic features of CT images. Front Oncol. 2023 Feb 13;13:1104316.

31. Min J, Dong F, Wu P, Xu X, Wu Y, Tan Y, et al. A Radiomic Approach to Access Tumor Immune Status by CD8+TRMs on Surgically Resected Non-Small-Cell Lung Cancer. OncoTargets Ther. 2021 Sep;Volume 14:4921–31.

32. de Sousa VML, Carvalho L. Heterogeneity in Lung Cancer. Pathobiology. 2018;85(1–2):96–107.

33. Yan X, Jiao SC, Zhang GQ, Guan Y, Wang JL. Tumor-associated immune factors are associated with recurrence and metastasis in non-small cell lung cancer. Cancer Gene Ther. 2017 Feb;24(2):57.

34. Zhu JY, Park T, Isola P, Efros AA. Unpaired Image-to-Image Translation using Cycle-Consistent Adversarial Networks [Internet]. arXiv; 2020 [cited 2023 Nov 13]. Available from: http://arxiv.org/abs/1703.10593

